# Genetic risk factor clustering within and across neurodegenerative diseases

**DOI:** 10.1101/2022.12.01.22282945

**Authors:** Mathew J. Koretsky, Chelsea Alvarado, Mary B Makarious, Dan Vitale, Kristin Levine, Sara Bandres-Ciga, Anant Dadu, Sonja W. Scholz, Lana Sargent, Faraz Faghri, Hirotaka Iwaki, Cornelis Blauwendraat, Andrew Singleton, Mike Nalls, Hampton Leonard

**Affiliations:** Center for Alzheimer’s Disease and Related Dementias, National Institutes of Health, Bethesda, MD, USA 20892; Laboratory of Neurogenetics, National Institute on Aging, National Institutes of Health, Bethesda, MD, USA 20892; Data Tecnica International LLC, Washington, DC USA 20037; University College London, London, UK; University of Illinois at Urbana-Champaign, Champaign, IL, 61820, USA; Neurodegenerative Diseases Research Unit, National Institute of Neurological Disorders and Stroke, Bethesda, MD, USA 20892; Department of Neurology, Johns Hopkins University, Baltimore, MD 21287; DZNE, Tuebingen, Germany

**Keywords:** Dementia, Single-nucleotide polymorphism, Genome-wide association study, Unsupervised, Machine learning

## Abstract

Overlapping symptoms and copathologies are common in closely related neurodegenerative diseases (NDDs). Investigating genetic risk variants across these NDDs can give further insight into disease manifestations. In this study we have leveraged genome-wide single nucleotide polymorphisms (SNPs) and genome-wide association study (GWAS) summary statistics to cluster patients based on their genetic status across identified risk variants for five NDDs (Alzheimer’s disease [AD], Parkinson’s disease [PD], amyotrophic lateral sclerosis [ALS], Lewy body dementia [LBD], and frontotemporal dementia [FTD]). The multi-disease and disease-specific clustering results presented here provide evidence that NDDs have more overlapping genetic etiology than previously expected and how neurodegeneration should be viewed as a spectrum of symptomology. These clustering analyses also show potential subsets of patients with these diseases that are significantly depleted for any known common genetic risk factors suggesting environmental or other factors at work. Establishing that NDDs with overlapping pathologies share genetic risk loci, future research into how these variants might have different effects on downstream protein expression, pathology and NDD manifestation in general is important for refining and treating NDDs.

## Introduction

Neurodegenerative diseases (NDDs), such as Alzheimer’s disease (AD), Parkinson’s disease (PD), amyotrophic lateral sclerosis (ALS), Lewy body dementia (LBD), and frontotemporal dementia (FTD), collectively affect more than 40 million people worldwide.^1^ This number is only expected to grow due to their mid-to late-life onset combined with an aging population.^1^ Genome-wide association studies (GWAS) have been used to identify common genetic variants linked to a higher risk of developing certain NDDs to uncover pathways that can lead to more advanced and targeted treatments.^2, 3^

Established risk variants for one NDD may play a role in the genetic makeup of several others. Overlapping risk variants across NDDs, even when sub-genome-wide significant in GWAS studies of a specific disease, may give insights into how disease can manifest across the spectrum of symptoms and copathologies shared by multiple NDDs.^4, 5^ Evidence of pleiotropic effects have been described previously in *GRN* for AD, PD, LBD, and ALS, *GBA* for PD and LBD, and *C9orf72* for FTD and ALS.^6–8^ Mutations in *MAPT* and *APOE* have also been linked to a range of NDDs and pathologies.^9, 10^ Additionally, it is possible to investigate how high and low genetic risk may manifest within a single disease, partitioning individuals with an NDD into classes across a quantitative range of genetic influence.

Here, we used genome-wide single nucleotide polymorphism (SNP) and GWAS summary statistics data for five NDDs (AD, PD, ALS, LBD, and FTD) to cluster patients based on their genetic status across identified risk variants for each disease. The multi-disease clusters presented here establish data-driven evidence of potential shared disease etiology that may explain overlapping symptomatology on the molecular level.^11^ The single-disease cluster analysis supports the idea that NDD specific risk data can inform both genetically different subtypes within the same disease and identify patients that may have the disease due to environmental, epigenetic, or other risk factors. This work seeks to refine NDD phenotypes and could help to differentiate NDDs for diagnosis and clinical trial enrollment.

## Materials and methods

### Samples

Supplementary Fig. 1 summarizes the workflow and data used for this project. The samples were obtained from public domain whole-genome sequencing (WGS) cohorts across the aforementioned NDDs. Existing genotype calls from the WGS cohorts were used and no additional genotype calling was performed. AD samples were obtained from the Alzheimer’s Disease Sequencing Project (ADSP), Alzheimer’s Disease Neuroimaging Initiative (ADNI), Mayo RNAseq Study (MayoRNAseq), Mount Sinai Brain Bank (MSBB), and the Religious Orders Study/Memory and Aging Project (ROSMAP).^12–14^ PD samples were obtained from Accelerating Medicines Partnership Parkinson’s disease (AMP-PD).^15^ FTD, LBD, and ALS data were all obtained from DementiaSeq.^16^ The total number of subjects across all cohorts was 23,885, of which 13,190 were cases (Supplementary Table 1). Only samples of genetically-determined European ancestry were used. For this analysis, 1000 cases for each disease were randomly sampled to ensure even representation, resulting in a final sample size of 5000 cases (Supplementary Table 1). GWAS summary statistics were obtained for each disease for use in the final SNP selection. The GWAS summary-level data used include Schwartzentruber et al. 2021 (AD), Nalls et al. 2019 (PD), Nicolas et al. 2018 (ALS), Chia et al. 2021 (LBD), and Ferrari et al. 2014 (FTD).^17–21^

### Genetic data quality control

Data from cohorts were not all on the same build; thus, data from cohorts using the hg19 build were lifted over to hg38.^22^ Summary statistics were lifted over as needed. Quality control (QC) was performed at both the individual cohort and combined cohort levels with Global Parkinson’s Genetics Program (GP2) pipelines (https://github.com/GP2code/) using PLINK (1.9 and 2). Sample level QC included genotype missingness (<0.02) as well as a duplicate removal (genetic relatedness matrix [GRM] cutoff of 0.95) and first cousin or closer relatedness pruning (GRM cutoff of 0.125). ADNI, ROSMAP, and AMP-PD also underwent genetic sex confirmation due to the availability of data for the X chromosome. Variant level QC included call rate pruning (<0.05) and pruning SNPs with a minor allele frequency (MAF) less than 0.05 for exclusion (Supplementary Table 2). After common SNPs were identified across the cohorts (explained in the systematic review section), the merged genotype data underwent an additional duplicate and relatedness check. The merged data was then passed through an ancestry prediction and pruning method to ensure all samples were of European descent (Supplementary Table 3). Ancestry was defined using reference panels from the 1000 Genomes Project and an Ashkenazi Jewish Population.^23, 24^ 50 principal components (PCs) were fit on 39,302 overlapping SNPs between the reference panel and the merged data. A classifier that was previously trained on the reference panel PCs was applied to the merged data PCs which returned the predicted ancestry label of each individual in the merged data. Samples predicted as non-European were removed.

### Systematic review

Prior to clustering across the NDDs, we narrowed the number and scope of SNPs. 17 million autosomal SNPs sequenced across all cohorts were identified and used to merge the individual cohort data. Disease-specific GWAS data was then used to filter for SNPs that reached genome-wide significance (i.e., p < 5e-08) in any one of the relevant GWAS studies. Using this SNP set, the merged data underwent munging and additional population substructure adjustment in GenoML.^25, 26^ Munging consists of pruning provided genotype data for linkage disequilibrium (LD) by removing any highly correlated genotypes in the sample series (r2 > 0.3 within a sliding window of 1Mb as minimum exclusion criteria). LD clumping and pruning was performed at random and was therefore not biased towards associations from any one disease. The adjustment process removes the effect of population substructure, which is further described in Makarious et al, 2022.^26^ The process required creating principal component analysis (PCA) loadings using the 5000 downsampled cases. Unlinked genome-wide SNPs outside of GWAS regions of interest were used to generate the 10 PCA loadings that approximate population substructure. The resulting 10 PCA loadings were used as covariates and regressed against the final SNP candidates using ordinary least squares regression. The resulting residual minor allele dosages were then z-normalized and used as the final output for model training at clustering. This process limits the effect of European population substructure from the genotypes before the clustering analysis is performed, mirroring the way in which GWAS utilizes covariates for population substructure within ancestry groups to reduce genomic inflation.^26^ After munging, the final SNP set consisted of 338 GWAS significant and population substructure adjusted SNPs not in LD with each other.

### Statistical analyses

Supplementary Fig. 2 summarizes the dimensionality reduction and clustering analyses performed. To effectively visualize and cluster the adjusted SNPs, Unified Manifold Approximation and Projection (UMAP) was chosen for dimensionality reduction. UMAP is a non-linear approach that is widely used in the field of population genetics.^27^ Using UMAP, 338 SNPs were reduced to 3 dimensions for each of the 5000 cases. Unsupervised clustering of the individuals was performed on the reduced data using the Mean Shift algorithm, as it is a deterministic algorithm that does not require the number of clusters to be specified, unlike more popular approaches such as K-Means clustering that require an *a priori* number of clusters to be defined.^28^

UMAP employs two fine-tuning hyperparameters, a and b, that impact the resulting embedding more specifically than minimum distance and spread. UMAP is a flexible algorithm that can be used on many different types and sizes of data; fine-tuning these hyperparameters enhances the model adjustment to the SNP data. Performing a grid search of a and b values from 0.25 to 3, with a step size of 0.25, the UMAP to Mean Shift pipeline was fitted and applied on a 70:30 (training:testing) split. To determine the best combination of hyperparameters, logistic regression was utilized with cluster membership as the input to predict an individual’s disease status for each NDD (AD, PD, ALS, LBD, and FTD). The chosen evaluation metric was the average area under the receiving operating characteristic (ROC) curve (AUC) across the disease-cluster regressions. The hyperparameters with the highest average AUC across the 144 tested combinations were then identified and used throughout the analysis (a = 2.75, b = 0.75).

UMAP is a stochastic algorithm; different runs can produce different results despite the input data and hyperparameters being the same. This can cause Mean Shift to identify a variable number of clusters in different iterations. To investigate this phenomenon, the UMAP to Mean Shift analysis was run on 15 different 70:30 (training:testing) splits for 100 iterations (i.e., 1500 iterations total), recording the number of clusters identified and the sample counts per cluster. Across the different splits and iterations, Mean Shift consistently identifies the main cluster that contains the majority (>4000 out of 5000 individuals) of the samples (Supplementary Table 4). From there, we applied an iterative clustering approach. Tracking samples across iterations, any sample consistently grouped into the main cluster was identified and labeled as a member of cluster 0 (C0). Conversely, any sample that was never grouped into this main cluster was labeled as a member of cluster 2 (C2). All the remaining samples that were not always grouped into the main cluster, according to the UMAP embedding, were labeled as a member of cluster 1 (C1). This process effectively addresses the variability caused by the stochastic nature of UMAP while capturing the clustering information provided by Mean Shift. More information on the iterative clustering approach can be found in the Supplementary Methods.

A z-test for proportions was performed to determine which multi-disease clusters were significantly enriched with certain NDDs compared to others. Next logistic regressions were utilized to see how cluster membership relates to NDD status as a complement to the previously described enrichment analysis. Cluster memberships were regressed on the set of 338 adjusted SNPs to identify any potential SNPs associated with increased likelihood of membership in a particular cluster, in part as a positive control, for loci with established pleiotropic associations (*GBA*, *GRN*, *LRRK2*, *MAPT*, *C9orf72*, and *APOE*). The Shapley values of the SNP predictors were then calculated, which is a popular game theory approach that helps explain and interpret how important each feature in a model is to the prediction of the dependent variable, in our case, cluster membership.^29^ SNPs most important in determining cluster membership were tracked across the clustering iterations to ensure consistency and a phenome-wide association study (PheWAS) look up was performed to further clarify the biological impact of the clusters.^30^ Finally, for each sample in the dataset, a polygenic risk score (PRS) was calculated for all five NDDs with the beta values from the same GWAS summary statistics used to determine the SNP set. From there, logistic regression was run to see how well the disease-specific PRSs determine cluster membership. Note that the PRS were z-score normalized to simplify the interpretation of the logistic regression output.

The same UMAP hyperparameter combination was used to perform the dimensionality reduction for each NDD in the disease-specific cluster analysis. Similar to the multi-disease analysis, when the UMAP to Mean Shift pipeline was applied to the 1000 samples for each NDD separately, the main cluster that contained a majority (>500 out of 1000 individuals) was formed consistently across iterations and diseases. Therefore, the iterative clustering approach from the multi-disease analysis was once again utilized. To account for the increased variability that comes with running the pipeline on a reduced sample size, individuals were grouped into C0 or C2 (C1 if only 2 clusters were identified in a disease subset) if they were consistently inside or outside the main cluster for at least 12 of the 15 70:30 (training:testing) splits (Supplementary Methods). The previously calculated PRS were re-normalized using the mean and standard deviations from the set of 1000 samples for each NDD. A T-test was run to see where the PRSs differentiated significantly from the mean of 0 that they were standardized to within the NDD-specific clusters.

To validate the methodology used and compare results, the clustering analysis was rerun under four different conditions. Firstly, the iterative clustering approach was compared to agglomerative hierarchical clustering on the same set of 5000 cases. Similar to Mean Shift, this popular tree-based approach does not require an *a priori* number of clusters to be defined.^31^ Next, the iterative clustering analysis was run with the *APOE* locus removed to ensure the clusters are not directly driven by E4 status due to its importance in determining NDD and AD risk. Next, the analysis was performed on a downsampled set of 500 cases for each disease to test the robustness of the results to sample size. Finally, a set of 1000 controls were included in the analysis to see how results are affected by the presence of a negative control. Note that for the downsampled and control-included analyses, QC and processing was exactly the same for the 2500 and 6000 samples, respectively. In addition, linkage disequilibrium score regression (LDSC) was run on the GWAS summary statistics to compare the genetic overlap seen in the clustering results to a more traditional method.

### Data and code availability

As previously mentioned, all samples for this analysis were obtained from public domain WGS cohorts. A repository containing all code for processing and analysis is publicly available to facilitate replication (https://github.com/NIH-CARD/NDD_risk_variant_clustering). In addition, an interactive website has been developed where researchers can further explore the described cluster memberships and results (https://nih-card-ndd-risk-variant-clustering-app-25rr5g.streamlitapp.com/).

## Results

### Multi-disease clustering

Using the iterative clustering approach, C0 contained 2,863 samples, C1 contained 2,074 samples, and C2 contained 63 samples (Fig. 1A, Supplementary Fig. 3). Regressions of each disease status per sample against cluster membership revealed that C0 was most significantly enriched with ALS (odds ratio [OR] = 1.631, p = 4.66e-08, beta = 0.489, standard error [SE] = 0.090), C1 with AD (OR 1.637, p = 9.20e-09, beta 0.493, SE 0.086), and C2 with FTD (OR = 3.063, p = 6.50e-05, beta = 1.119, SE = 0.280). C2 was enriched with PD compared to the overall disease distributions but not to the point of significance in the regressions (Table 1). After multiple test corrections, none of the clusters were significantly enriched with LBD.

**Figure 1.**
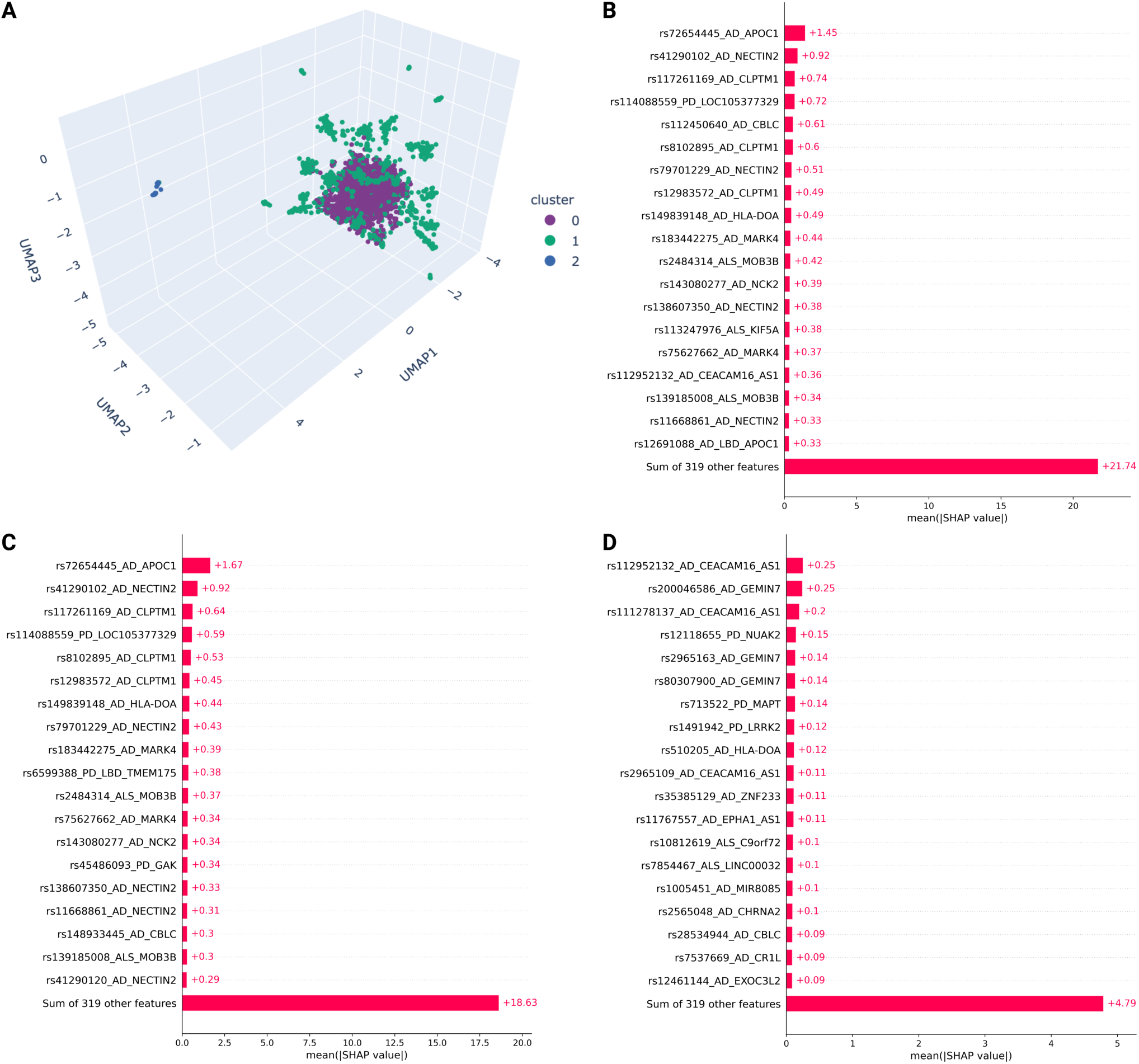
Multi-disease cluster membership. **(A)** Multi-disease clusters. Shapley values of SNPs most impacting the defining of **(B)** cluster 0, **(C)** cluster 1, and **(D)** cluster 2.

**Table 1.** Disease association summary statistics and frequency per multi-disease cluster.

| **Disease** | **Multi-disease cluster membership** | **OR** | **BETA** | **SE** | **P** | **% samples with disease** |
| --- | --- | --- | --- | --- | --- | --- |
| AD | Cluster 0 | 0.646 | -0.436 | 0.086 | 3.51e-07 | 0.165* |
|  | Cluster 1 | 1.637 | 0.493 | 0.086 | 9.20e-09 | 0.252* |
|  | Cluster 2 | 0.245 | -1.408 | 0.596 | 0.018 | 0.079 |
| PD | Cluster 0 | 1.141 | 0.132 | 0.086 | 0.125 | 0.210 |
|  | Cluster 1 | 0.859 | -0.152 | 0.087 | 0.080 | 0.186 |
|  | Cluster 2 | 1.308 | 0.269 | 0.322 | 0.404 | 0.238* |
| ALS | Cluster 0 | 1.631 | 0.489 | 0.090 | 4.66e-08 | 0.226* |
|  | Cluster 1 | 0.646 | -0.437 | 0.090 | 1.15e-06 | 0.167* |
|  | Cluster 2 | 0.245 | -1.408 | 0.596 | 0.018 | 0.079* |
| LBD | Cluster 0 | 0.836 | -0.179 | 0.084 | 0.033 | 0.190 |
|  | Cluster 1 | 1.198 | 0.180 | 0.084 | 0.032 | 0.213 |
|  | Cluster 2 | 1.004 | 4.30e-03 | 0.341 | 0.990 | 0.206 |
| FTD | Cluster 0 | 1.021 | 0.021 | 0.085 | 0.802 | 0.208 |
|  | Cluster 1 | 0.895 | -0.111 | 0.086 | 0.196 | 0.182* |
|  | Cluster 2 | 3.063 | 1.119 | 0.280 | 6.50e-05 | 0.397* |

\*denotes a p-value < 0.05 for the frequency increase or decrease in a certain disease status per cluster compared to the null estimate of 20%.

The PRS regressions revealed that the only PRS that was significantly associated with membership in all clusters was AD. C0 and C2 had negative associations (defining non-AD driven clusters), while C1 had a positive association. The trend of C0 having significant negative associations and C1 having significant positive associations continued for the PD, ALS, and LBD PRS, while no other PRS was shown to be significantly associated with C2. The FTD PRS was not significantly associated with membership in any of the clusters. For a summary of these results, please refer to Table 2. The PRS distributions by cluster are displayed in Fig. 2.

**Figure 2.**
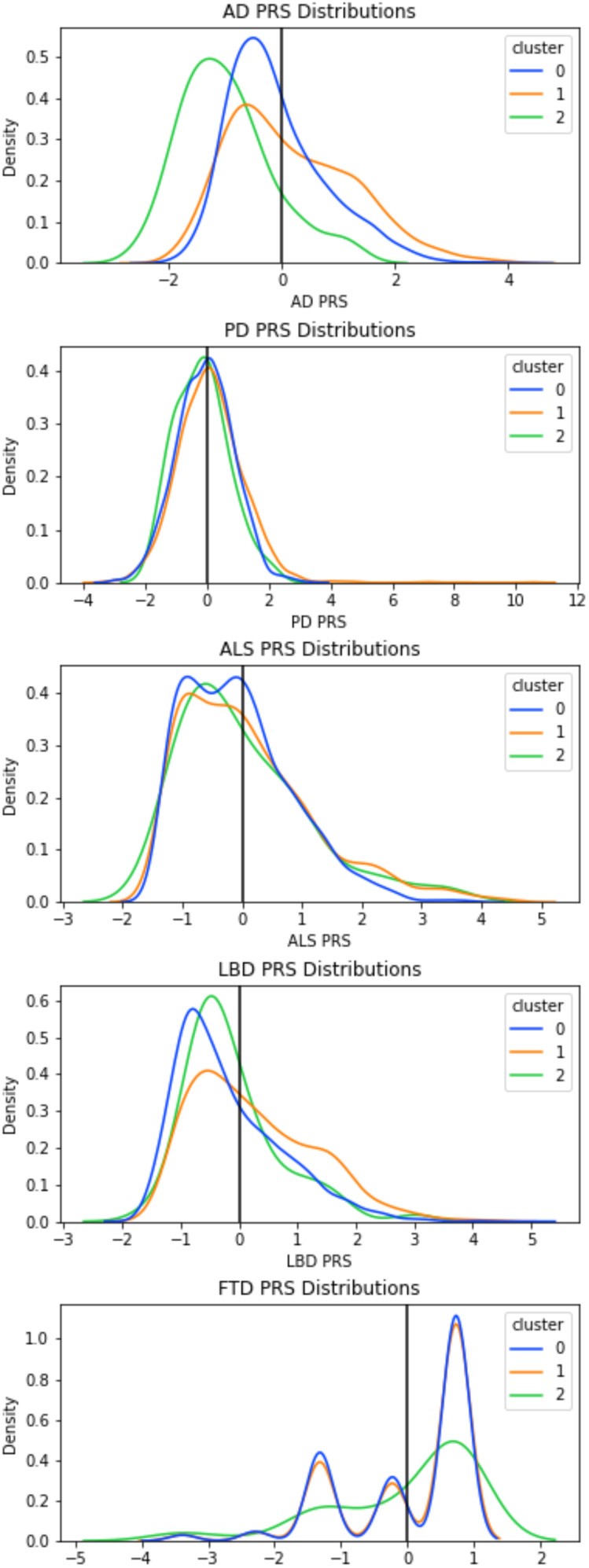
Standardized PRS distributions per multi-disease cluster.

**Table 2.** PRS association summary statistics per cluster for both the multi-disease and single-disease clustering analyses Disease.

| Disease<br>/ PRS | Multi-disease clustering |  |  |  |  | Disease-specific clustering |  |  |  |  |
| --- | --- | --- | --- | --- | --- | --- | --- | --- | --- | --- |
|  | Cluster | OR | BETA | SE | P | Cluster | OR | BETA | SE | P |
| AD | Cluster 0 | 0.804 | -0.218 | 0.034 | 2.31e-10 | Cluster 0 | 0.918 | -0.086 | 0.077 | 0.266 |
|  | Cluster 1 | 1.331 | 0.286 | 0.035 | 2.32e-16 | Cluster 1 | 1.117 | 0.111 | 0.078 | 0.154 |
|  | Cluster 2 | 0.146 | -1.921 | 0.249 | 1.31e-14 | Cluster 2 | 0.550 | -0.598 | 0.425 | 0.160 |
| PD | Cluster 0 | 0.838 | -0.177 | 0.035 | 3.95e-07 | Cluster 0 | 0.826 | -0.192 | 0.080 | 0.017 |
|  | Cluster 1 | 1.209 | 0.190 | 0.035 | 6.38e-08 | Cluster 1 | 1.211 | 0.192 | 0.080 | 0.017 |
|  | Cluster 2 | 0.829 | -0.187 | 0.144 | 0.192 |  |  |  |  |  |
| ALS | Cluster 0 | 0.858 | -0.154 | 0.034 | 7.00e-06 | Cluster 0 | 0.577 | -0.549 | 0.093 | 3.69e-09 |
|  | Cluster 1 | 1.171 | 0.158 | 0.034 | 4.00e-06 | Cluster 1 | 1.732 | 0.549 | 0.093 | 3.69e-09 |
|  | Cluster 2 | 0.950 | -0.051 | 0.142 | 0.721 |  |  |  |  |  |
| LBD | Cluster 0 | 0.653 | -0.427 | 0.036 | 1.31e-32 | Cluster 0 | 0.627 | -0.466 | 0.081 | 7.36e-09 |
|  | Cluster 1 | 1.549 | 0.437 | 0.036 | 5.22e-34 | Cluster 1 | 1.594 | 0.466 | 0.081 | 7.36e-09 |
|  | Cluster 2 | 0.872 | -0.137 | 0.148 | 0.354 |  |  |  |  |  |
| FTD | Cluster 0 | 0.977 | -0.023 | 0.034 | 0.498 | Cluster 0 | 0.988 | -0.012 | 0.077 | 0.872 |
|  | Cluster 1 | 1.029 | 0.029 | 0.034 | 0.402 | Cluster 1 | 1.039 | 0.038 | 0.078 | 0.625 |
|  | Cluster 2 | 0.920 | -0.084 | 0.132 | 0.528 | Cluster 2 | 0.777 | -0.252 | 0.232 | 0.278 |

Based on the Shapley values for individual SNPs, important variants determining membership in C0 and C1 were localized to *APOC1* (rs72654445) and *CEACAM16*/*AS1* (rs112952132 and rs111278137) for C2 (Fig. 1B-D). Two SNPs in *NECTIN2* (rs41290102 and rs79701229) were of high importance for differentiating C0 and C1 from C2. All specified variants, aside from rs41290102, show significant associations with low density lipoprotein cholesterol levels as well as high cholesterol in the Open Targets PheWAS look up (Supplementary Table 5). Variants belonging to the genes *APOC1*, *CBLC*, *CEACAM16/AS1*, *CLPTM1*, and *NECTIN2* were identified as significant drivers in at least one multi-disease cluster based on their mean absolute Shapley value. All these top variants are associated with AD and cluster within 1Mb of the *APOE* locus on chromosome 19 and likely driven by a connection to variable E4 allele risk.^32^ Variants associated with the specified loci only account for 20.99%, 20.97%, and 8.90% of the differentiation between C0, C1, and C2, respectively. In addition, of the top 20 variants associated with each cluster, 13 were consistent across clustering iterations for C0 and C1, and 9 were consistent for C2 (Supplementary Table 6).

The individual SNP regressions reveal variants localized with *APOE*, *GBA*, and *LRRK2* are significantly associated with all clusters (Supplementary Table 7). Additionally, variants localized to *MAPT* (rs713522) and *C9orf72* (rs17696570) showed significant associations with C0 and C1. The *GBA* variant (rs76763715) shows a strong positive association with C1 (OR = 8.020, p = 0.008, Beta = 2.082, SE = 0.782) and a very strong negative association with C2 (OR = 8.33e-04, p = 0.023, Beta = −7.091, SE = 3.110). One of two variants that determine *APOE4* status was in the set of 338 SNPs used for clustering (rs7412). Given its importance in determining NDD and AD risk, it should be noted that the variant itself is not significantly associated with membership in any of the clusters (Supplementary Table 8).

### Disease-specific clustering

For the NDD-specific clusters, only AD and FTD had a group of samples that were consistently outside the main cluster across all iterations (i.e., a presence of C2), containing 14 and 25 samples, respectively (Supplementary Fig. 4). Interestingly, across all single-disease clustering analyses, the AD PRS is significantly associated with differentiating subsets of samples (Table 2 and Supplementary Table 9).

In the AD-specific analysis, C1 showed significant genetic risk enrichment for AD. This same cluster also demonstrated significant enrichment of ALS and LBD genetic risk factors. In the PD-specific analysis, C0 was significantly enriched for AD genetic risk, and C1 for LBD genetic risk. In ALS, there are two clearly defined clusters, one significantly enriched for ALS genetic risk and the other depleted for genetic risk. The ALS cluster that has increased ALS genetic risk shows a significant decrease in AD genetic risk loading. For LBD, C0 shows a significant decrease in both AD and LBD genetic risk. In contrast, C1 shows a significant increase in AD and LBD genetic risk, suggesting a less genetically influenced form of the disease is likely. FTD disease-specific clusters were complicated, as the FTD genetic risk was not significantly associated with any of the three clusters identified. What is of interest is that one FTD cluster is significantly enriched for AD, PD, ALS, and LBD genetic risk, while another cluster is significantly depleted with regard to genetic risk for PD, ALS, and LBD.

### Comparative analyses

Supplementary Table 10 compares the multi-disease cluster counts from the iterative clustering approach with the four conditional analyses described previously. Hierarchical clustering also identifies three clusters that show a similar pattern to the iterative clustering approach (Supplementary Fig. 5). Supplementary Fig. 6 contains a dendrogram to visualize the hierarchical relationship between the clusters formed. The hierarchical clusters reveal a much larger C0 (4,073 cases compared to 2,863 cases) and a much smaller C1 (857 cases compared to 2,074 cases). However, when looking at the disease enrichments per cluster the only changes in significance are the presence of PD in C1 and ALS in C2 (Supplementary Table 11).

The proportion of samples in C2 increased for the analysis with the *APOE* locus removed when compared to the original analysis (Supplementary Fig. 7). Despite this, the regression of disease status against cluster membership shows C0 is still most enriched with ALS (OR = 1.752, p = 2.3E-08, Beta = 0.561, SE = 0.090), C1 with AD (OR = 1.701, p = 6.1E-10, Beta = 0.531, SE = 0.086), and C2 with FTD (OR = 1.855, p = 0.021, Beta = 0.618, SE = 0.269). C2 is significantly enriched with LBD, but not to the point of significance in the regressions, revealing that the increase in C2 size is largely due to LBD cases (Supplementary Table 12). The PRS regressions revealed a similar trend, with PRS for AD, PD, ALS, and LBD having significant negative associations with membership in C0 and significant positive associations with membership in C1 (Supplementary Table 13). This shows a similar pattern to the original analysis and a distinct group of samples that are consistently depleted for genetic risk.

The proportion of samples in each cluster is very similar for the downsampled analysis when compared to the 5000 sample analysis (Supplementary Fig. 8). The regression of disease status against cluster membership once again shows C0 is most enriched with ALS (OR = 1.731, p = 1.5E-05, Beta = 0.549, SE = 0.127), C1 with AD (OR = 1.735, p = 6.0E-06, Beta = 0.551, SE = 0.121), and C2 with FTD (OR = 2.351, p = 0.042, Beta = 0.855, SE = 0.421). Again, none of the clusters show significant enrichment of LBD (Supplementary Table 14). The same trend in PRS regressions was observed, with PRS for AD, PD, ALS, and LBD having significant negative associations with membership in C0 and significant positive associations with membership in C1 (Supplementary Table 15).

Of the 1000 controls included as a negative control, 991 are grouped into C0 (Supplementary Fig. 9). Because of this C0 is most significantly enriched with controls (OR = 108.690, p = 6.1E-25, Beta = 4.688, SE = 0.450) and is significantly depleted for all NDDs except ALS (Supplementary Table 16). It continues to be the case that C1 is most enriched with AD (OR = 2.097, p = 1.5E-18, Beta = 0.740, SE = 0.084) and C2 with FTD (OR = 108.690, p = 6.1E-25, Beta = 4.688, SE = 0.450). Once again, C0 shows significant negative associations with PRS for each NDD besides FTD, which is expected as nearly every control is contained within the cluster (Supplementary Table 17).

The LDSC analysis showed that AD and PD (r = 0.197, SE = 0.084, p = 0.019), AD and LBD (r = 0.385, SE = 0.188, p = 0.040), and PD and LBD (r = 0.599, SE = 0.166, p = 3.0E-04) were the only pairs of NDDs to have significant genetic correlations (Supplementary Table 18). This is less genetic overlap than is seen in the NDD-specific clustering analysis, where significant associations were seen between the AD PRS and cluster membership in all studied NDDs, as well as the ALS PRS and cluster membership for AD and FTD (Supplementary Table 9).

## Discussion

### Utility in disease subtyping and clinical trials

The clustering results support the idea that closely related NDDs have more overlapping genetic etiology than previously expected, using data-driven approaches to show how in many cases, neurodegeneration should be viewed as a spectrum of symptomology and risk factors, not discrete units. This is evident in that we have shown that all three of the multi-disease clusters have members from each studied NDD, and each of these clusters shows a significant association with variants from loci that have been previously linked to multiple NDDs. We have also shown these clusters to be robust to changes in SNP selection, sample size, the inclusion of the *APOE* locus, and the presence of controls. Synthesizing the results of our disease-specific and multi-disease clustering, we note that major differentiating factors between patients seem to be a general lack of genetic risk or a mix of disease risk enrichments at varying degrees (corresponding to cluster 0). This exemplifies the need for future studies of environmental and epigenetic risk factors shared across NDDs. We also suggest that repeating these analyses in a large set of harmonized pathology derived data would provide downstream insights on shared mechanisms in the brains of affected patients within and across diseases.

The overlapping deviations between the disease-specific clusters and the PRS for various NDDs provide evidence that neurodegeneration lies on a spectrum.^33^ While it is possible our participant-level analysis may be more optimistic compared to summary statistics-based methods, these overlaps show there are more genetic associations between NDDs than previously revealed through traditional methods such as LDSC. There may be groups of patients diagnosed with one NDD but have a high genetic risk for another. For example, the PRS for LBD, a disease that is already known to be closely related to PD and AD in terms of both clinical and pathological manifestations, has significant associations with cluster membership in all of the other presented NDDs.^34–36^ Overlaps like these show the need for further research into refining phenotypes for the diagnosis of NDDs, as well as closer monitoring of individuals post-diagnosis to see if changes occur that may cause the need for reconsideration of treatments. Understanding that NDDs with overlapping pathologies tend to share genetic risk loci, in diagnosis and clinical trial enrollment, it will be important to determine how the variants are most strongly associated with each NDD. More importantly, understanding how the variants might have subtly different effects on downstream protein expression (i.e., tau pathology, alpha-synuclein expression) and pathology that influence disease manifestation will be valuable for precision clinical trials.

### Limitations

The limitations of this research include a lack of diversity, insufficient clinical data across sample series, case imbalances between diseases that limited total sample sizes, a lack of rare variant inclusion, and a lack of information on insertions, deletions, and expansions.^37^ In particular, the FTD PRS estimates suffered from the small GWAS sample size and that may have impacted the results. Given the limited availability of non-European samples, it is difficult to appropriately model any effects ancestral differences may introduce. Similarly, the imbalance between disease sample sizes was a significant limitation. The imbalance resulted in the use of 1000 cases from each disease. The decision to use 1000 cases from each disease resulted from the ALS, the smallest cohort, only having 1105 cases. The clustering model would benefit from having more samples in order to make it more robust to outliers and to potentially identify any other potential clusters not captured in the currently sampled cohorts. The number of cases per disease could have been increased to 2000 if ALS and FTD were removed from the analysis, however the scope of the results would have been limited since LBD has previously been shown to be closely related to AD and PD both clinically and pathologically. In addition, we have shown the results to be robust to downsampling to 500 cases per disease, which mitigates some concern about the sample size used in the clustering analysis. The lack of broad and uniform clinical/phenotypic data across cohorts limited analyses and translational conclusions. The only common phenotype data common across cohorts was sex and European ancestry. Age collection across cohorts varied with no common collection point (i.e., age at onset, age at death, etc.) and differing age measures from precise ages to age range bins. Additionally, the quality of phenotypes and the impact of “proxy-cases” or self-reported cases in some large biobank studies may impact the overlap across diseases as, ideally, all phenotypes would be corroborated by imaging or pathology. Other clinical traits that would have been useful for further analyses include family history, disease severity, and medication status. The lack of rare variant inclusion implies that the clustering model may not identify acute genetic differences between NDDs. This lends to these clusters being quite broad and focused on sporadic manifestations of these NDDs, likely not establishing contrasts that could be attributed to early-onset familial cases. Lastly, because the clusters formed here are based on individual SNPs they do not account for insertions, deletions, or expansions. For example, pathology overlaps are seen between ALS and FTD cases due to repeat expansions in *C9orf72.*^8^ This limits the amount of genetic overlap between diseases that can be captured by the clusters.

## Conclusion

This report used data driven methods to define the spectrum of neurodegenerative disease interconnectivity. These connections between diseases are based on shared genetic risk factors and the interplay between these risk factors, often recapitulated in symptomology and pathology. Using this data, we can better understand potential fine-grained diagnoses that incorporate more variability than previous discrete classifications of neurodegenerative diseases.

## Abbreviations

AD: Alzheimer’s disease
ADNI: Alzheimer’s disease Neuroimaging Initiative
ADSP: Alzheimer’s Disease Sequencing Project
ALS: Amyotrophic lateral sclerosis
AMP-PD: Accelerating Medicines Partnership Parkinson’s disease
AUC: Area under the receiving operating characteristic curve
FTD: Frontotemporal dementia
GP2: Global Parkinson’s Genetics Program
GRM: Genetic relatedness matrix
GWAS: Genome-wide association study
LBD: Lewy body dementia
LD: Linkage disequilibrium
MAF: Minor allele frequency
MayoRNAseq: Mayo RNAseq Study
NDD: Neurodegenerative disease
OR: Odds ratio
PC: Principal component
PCA: Principal component analysis
PD: Parkinson’s disease
PRS: Polygenic risk score
QC: Quality control
ROC: Receiving operating characteristic
ROSMAP: Religious Orders Study/Memory and Aging Project
SE: Standard error
SNP: Single nucleotide polymorphism
UMAP: Unified Manifold Approximation and Projection
WGS: Whole genome sequencing

## Data Availability

All samples for this analysis were obtained from public domain WGS cohorts. A repository containing all code for processing and analysis is publicly available to facilitate replication (https://github.com/NIH-CARD/NDD_risk_variant_clustering). In addition, an interactive website has been developed where researchers can further explore the described cluster memberships and results (https://nih-card-ndd-risk-variant-clustering-app-25rr5g.streamlitapp.com/).

## Acknowledgements

This research was supported in part by the Intramural Research Program of the NIH, National Institute on Aging (NIA), National Institutes of Health, Department of Health and Human Services; project number ZO1 AG000535, as well as the National Institute of Neurological Disorders and Stroke (1ZIANS003154).

Data used in the preparation of this article were obtained from the AMP-PD Knowledge Platform. For up-to-date information on the study, visit https://www.amp-pd.org. AMP-PD – a public-private partnership – is managed by the FNIH and funded by Celgene, GSK, the Michael J. Fox Foundation for Parkinson’s Research, the National Institute of Neurological Disorders and Stroke, Pfizer, and Verily. We would like to thank AMP-PD for the publicly available whole-genome sequencing data, including cohorts from the Fox Investigation for New Discovery of Biomarkers (BioFIND), the Parkinson’s Progression Markers Initiative (PPMI), and the Parkinson’s Disease Biomarkers Program (PDBP). The Parkinson’s Disease Biomarker Program (PDBP) consortium is supported by the National Institute of Neurological Disorders and Stroke (NINDS) at the National Institutes of Health. A full list of PDBP investigators can be found at https://pdbp.ninds.nih.gov/policy. Harvard Biomarker Study (HBS) is a collaboration of HBS investigators (full list of HBS investigators found at https://www.bwhparkinsoncenter.org/biobank) and funded through philanthropy and NIH and Non-NIH funding sources. The HBS Investigators have not participated in reviewing the data analysis or content of the manuscript. The DementiaSeq data were obtained from dbGap (accession number: phs001963.v2.p1).

The results published here are in whole or in part based on data obtained from the AD Knowledge Portal (https://adknowledgeportal.org). Data generation was supported by the following NIH grants: P30AG10161, P30AG72975, R01AG15819, R01AG17917, R01AG036836, U01AG46152, U01AG61356, U01AG046139, P50 AG016574, R01 AG032990, U01AG046139, R01AG018023, U01AG006576, U01AG006786, R01AG025711, R01AG017216, R01AG003949, R01NS080820, U24NS072026, P30AG19610, U01AG046170, RF1AG057440, and U24AG061340, and the Cure PSP, Mayo and Michael J Fox foundations, Arizona Department of Health Services and the Arizona Biomedical Research Commission. We thank the participants of the Religious Order Study and Memory and Aging projects for the generous donation, the Sun Health Research Institute Brain and Body Donation Program, the Mayo Clinic Brain Bank, and the Mount Sinai/JJ Peters VA Medical Center NIH Brain and Tissue Repository. Data and analysis contributing investigators include Nilüfer Ertekin-Taner, Steven Younkin (Mayo Clinic, Jacksonville, FL), Todd Golde (University of Florida), Nathan Price (Institute for Systems Biology), David Bennett, Christopher Gaiteri (Rush University), Philip De Jager (Columbia University), Bin Zhang, Eric Schadt, Michelle Ehrlich, Vahram Haroutunian, Sam Gandy (Icahn School of Medicine at Mount Sinai), Koichi Iijima (National Center for Geriatrics and Gerontology, Japan), Scott Noggle (New York Stem Cell Foundation), Lara Mangravite (Sage Bionetworks).

This study used the high-performance computational capabilities of the Biowulf Linux cluster at the National Institutes of Health (http://hpc.nih.gov).

## Supplementary material

Supplementary material is available at *Brain* online.

