## Supplementary Material for "Genetic risk factor clustering within and across neurodegenerative diseases"

- Supplementary Figures 1-9
- Supplementary Tables 1-18
- Supplementary Methods

**Supplementary Figure 1**

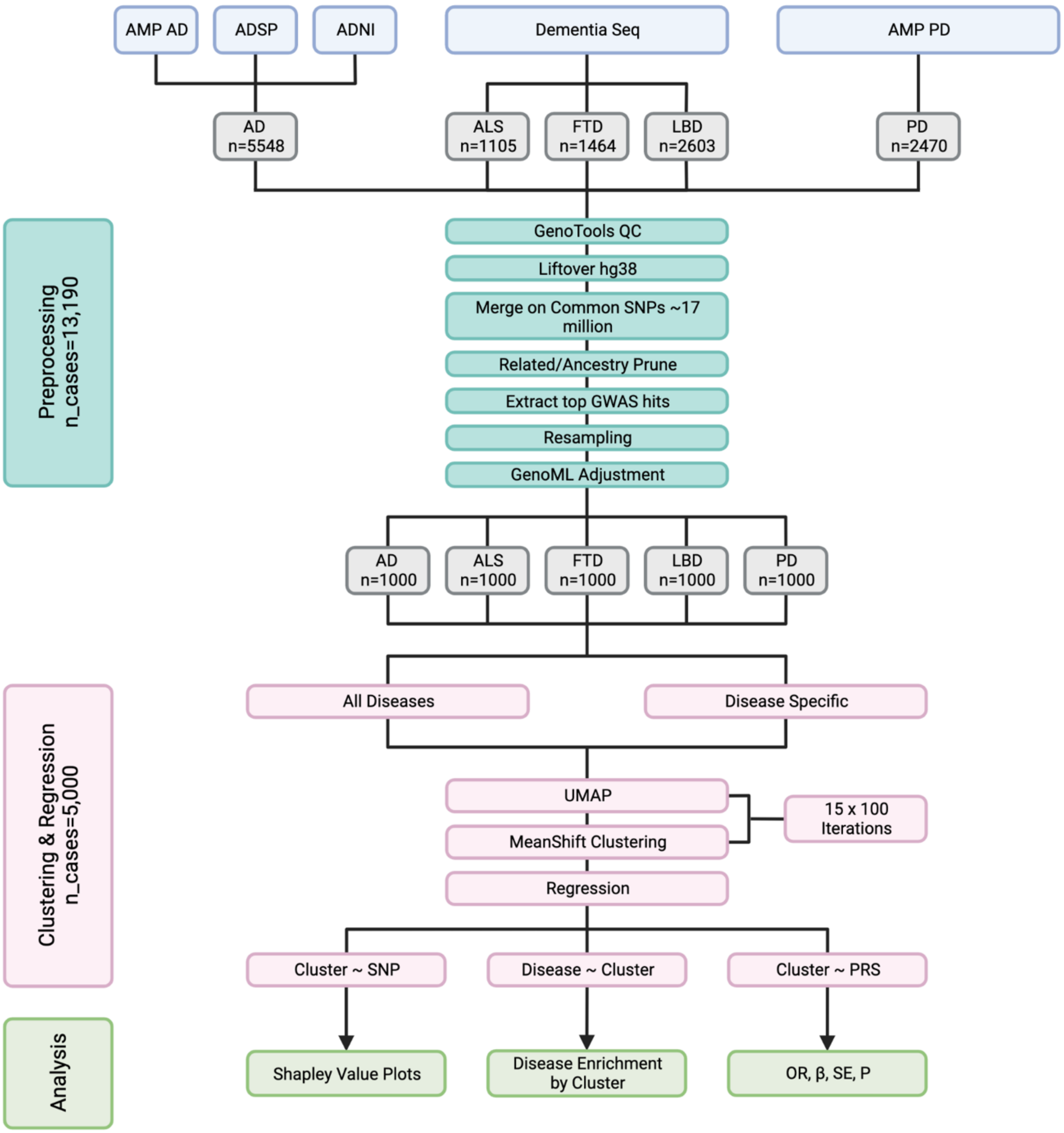


**Workflow diagram summarizing cohort information, preprocessing and statistical analysis performed.**

**Supplementary Figure 2**

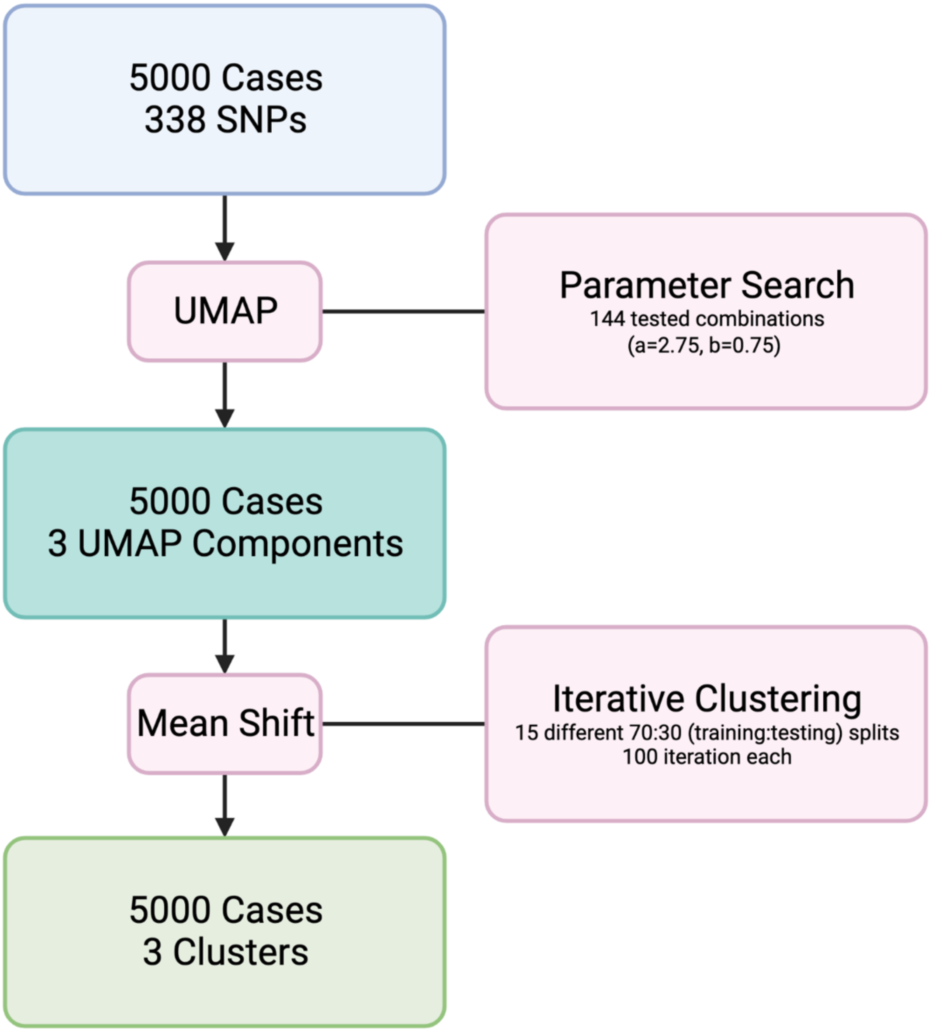


**Workflow diagram summarizing the dimensionality reduction and clustering analyses performed.**

**Supplementary Figure 3**

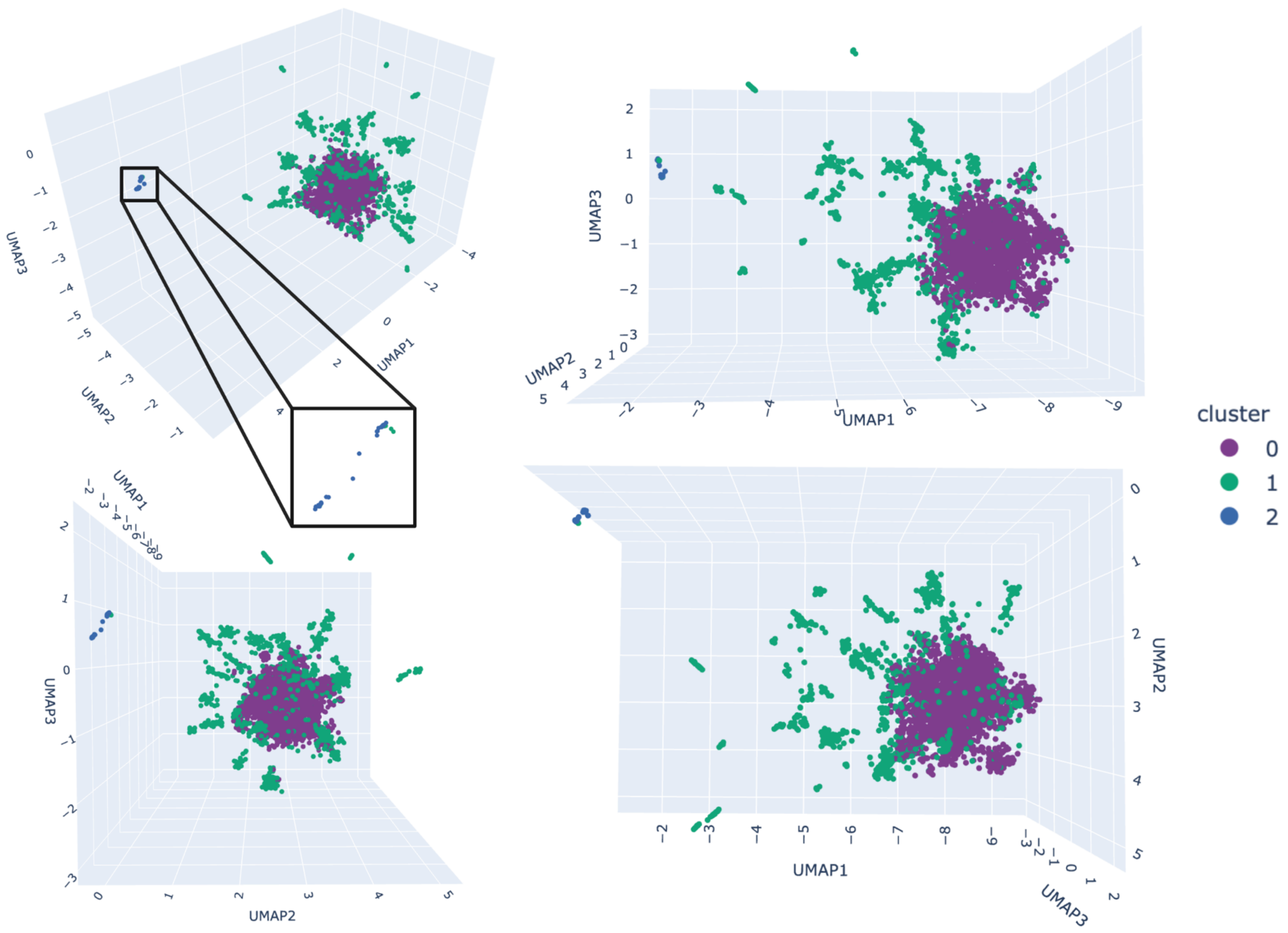


**Multi-disease clusters resulting from the iterative clustering approach.**

**Supplementary Figure 4**

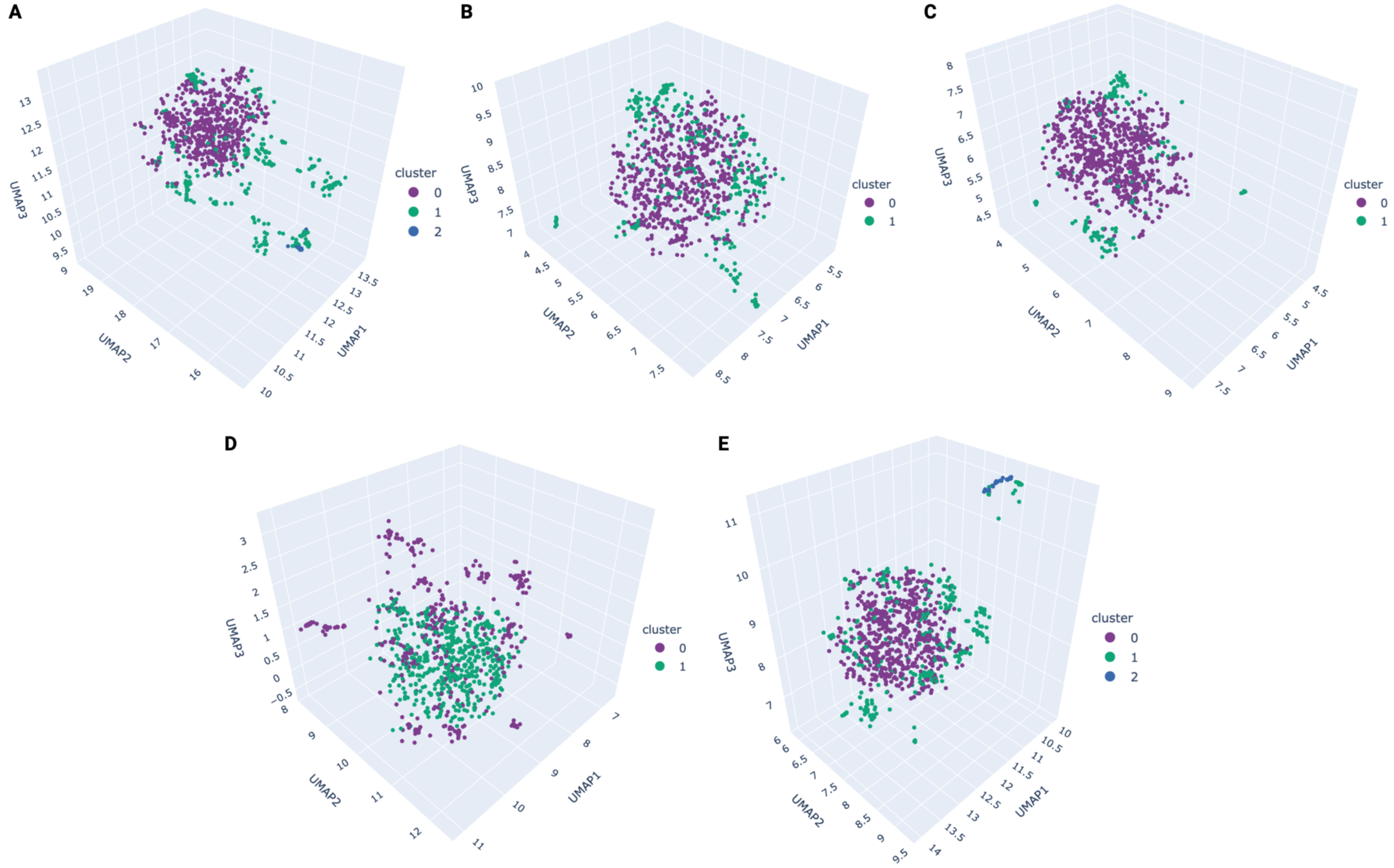


**Disease-specific cluster memberships. (A)** Alzheimer's disease (AD). **(B)** Parkinson's disease (PD). **(C)** Amyotrophic lateral sclerosis (ALS). **(D)** Lewy body dementia (LBD). **(E)** Frontotemporal dementia (FTD).

**Supplementary Figure 5**

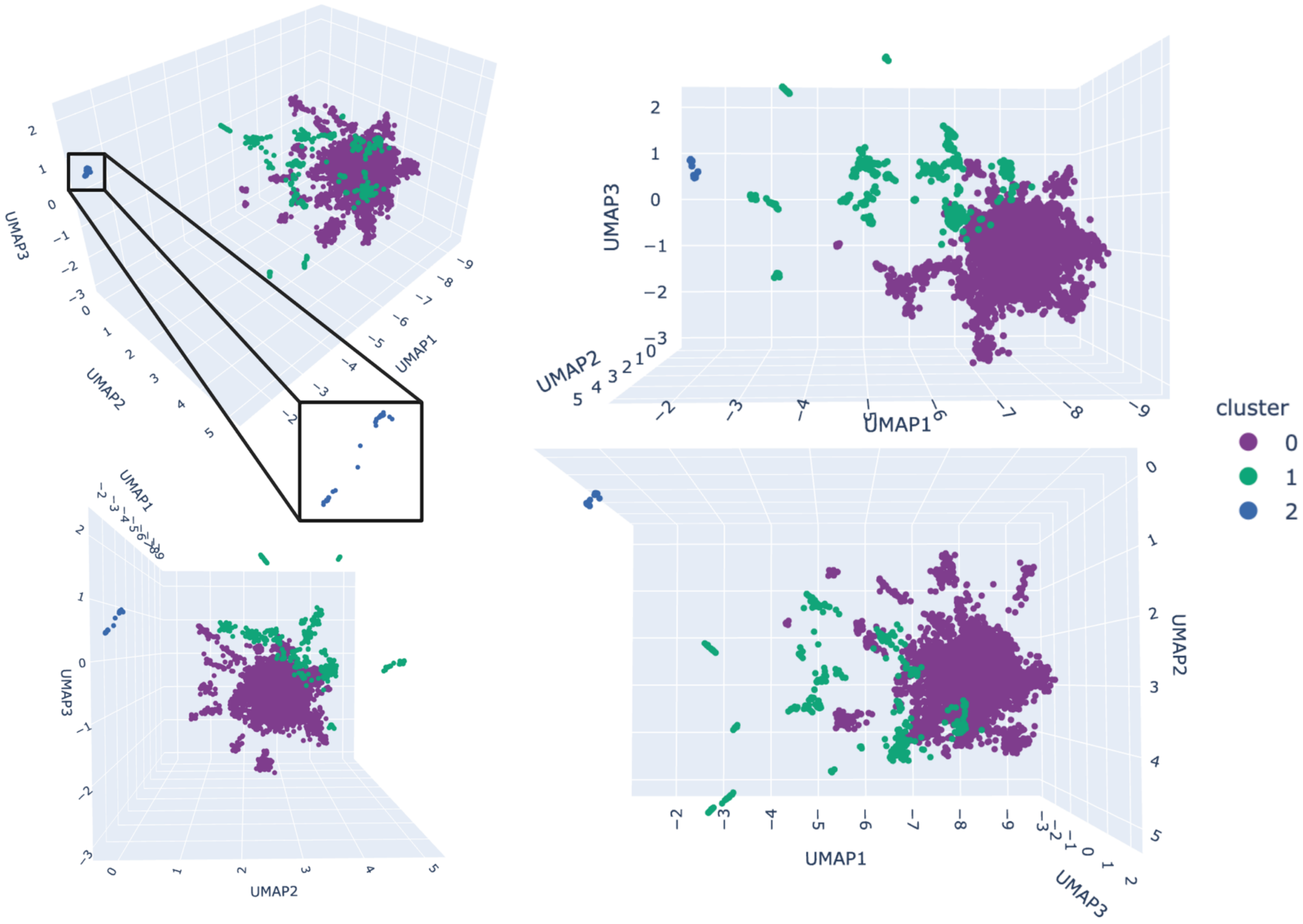


**Multi-disease clusters resulting from hierarchical clustering.**

**Supplementary Figure 6**

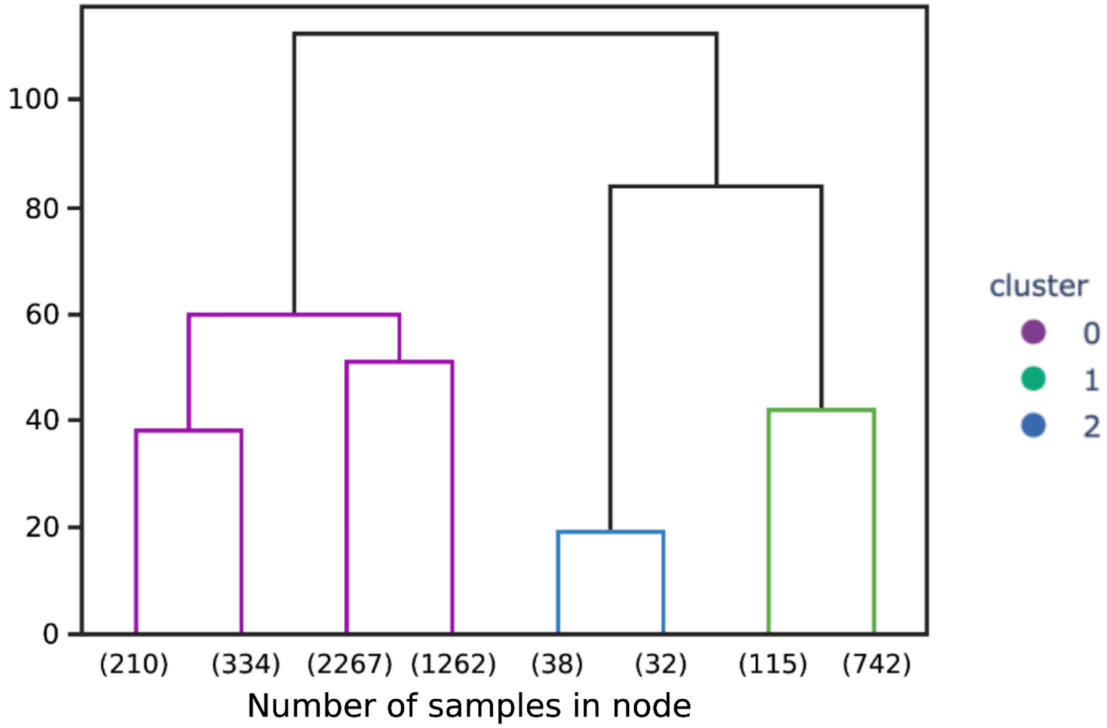


**Hierarchical clustering dendrogram.**

**Supplementary Figure 7**

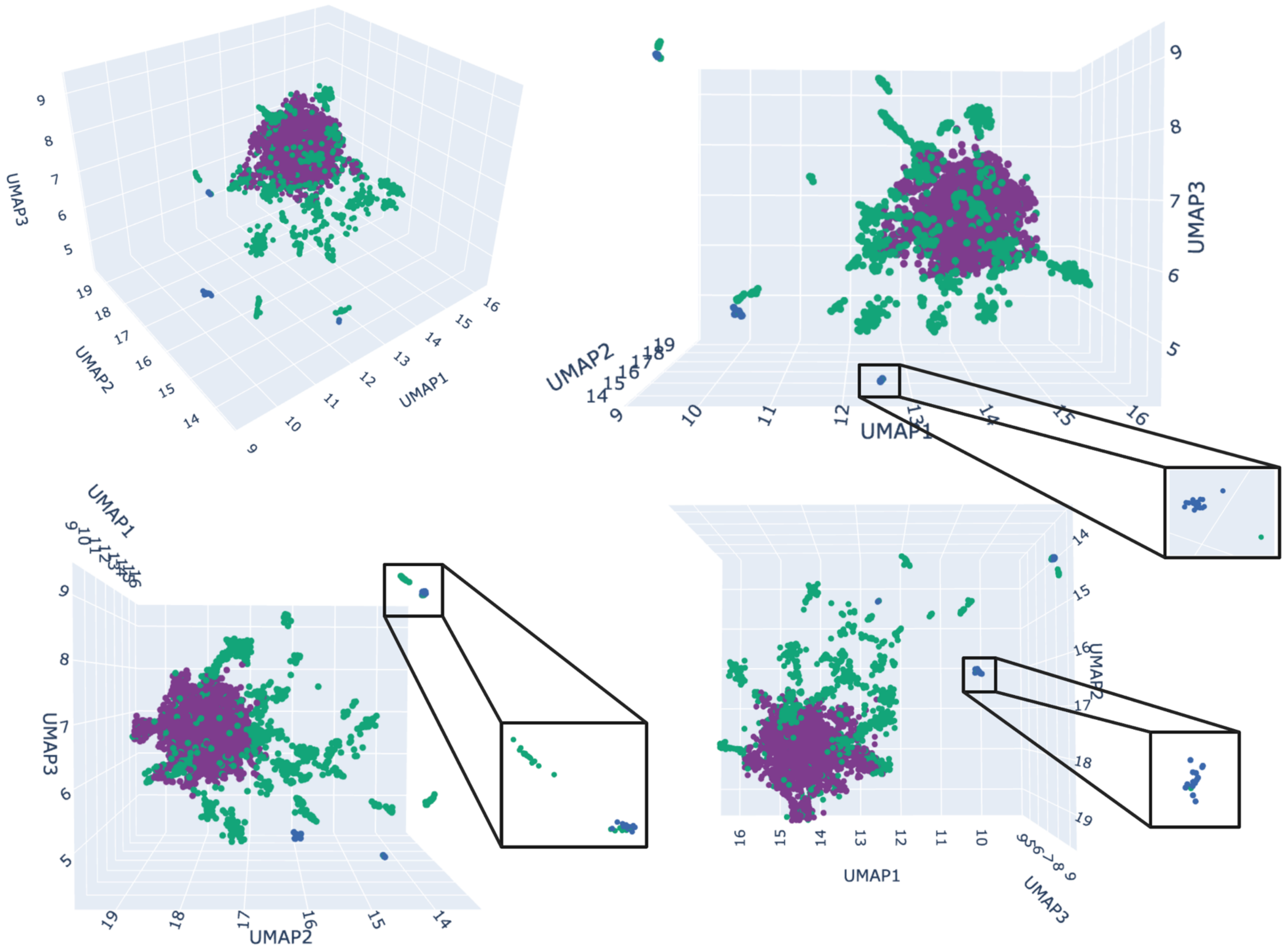


**Multi-disease clusters resulting from the iterative clustering approach with the *APOE* locus removed.**

**Supplementary Figure 8**

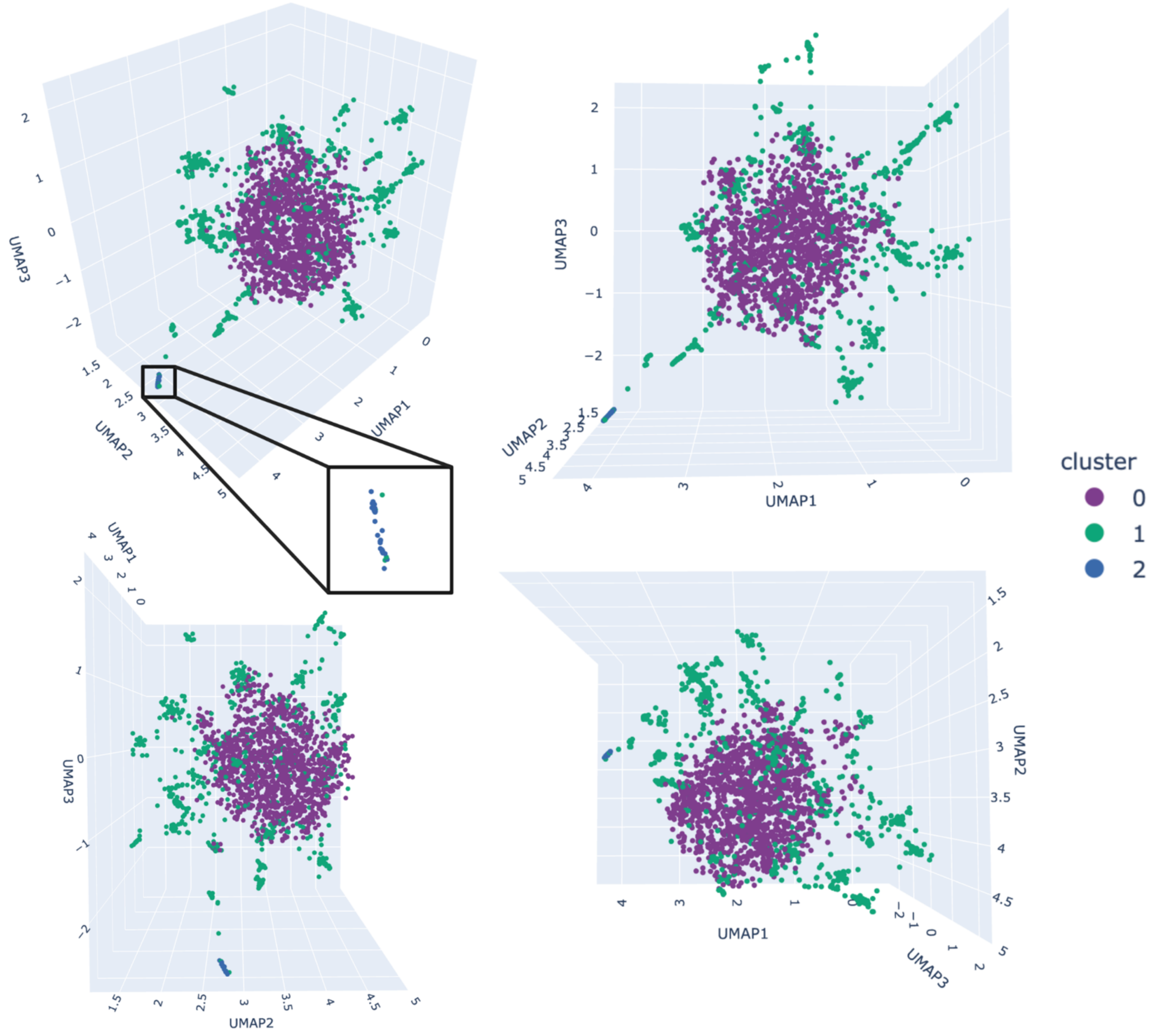


**Multi-disease clusters resulting from the iterative clustering approach with downsampled data (500 cases per disease).**

**Supplementary Figure 9**

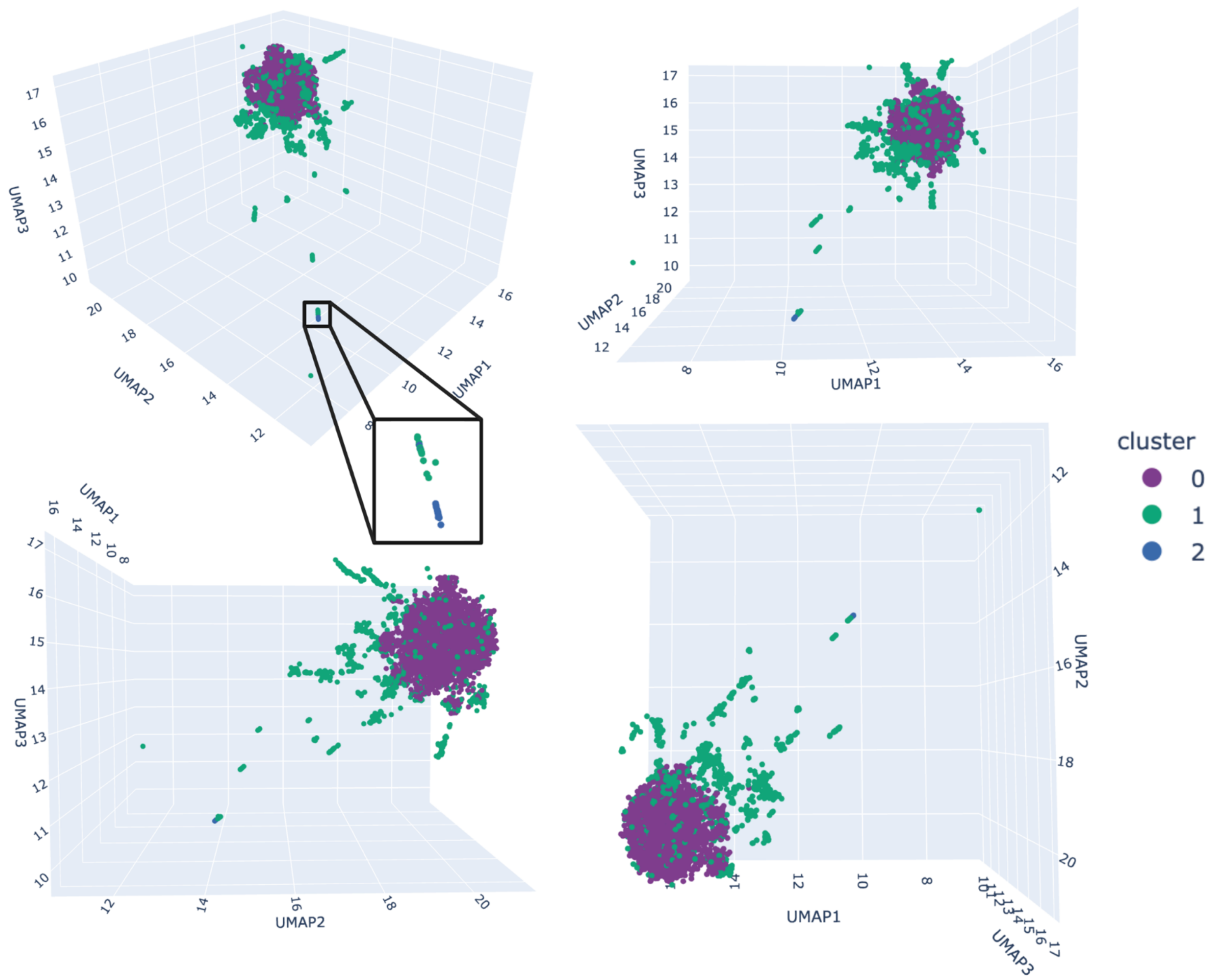


**Multi-disease clusters resulting from the iterative clustering approach with 1000 controls included as a negative control.**

**Supplementary Table 1 Cohort information**

| Cohort | Disease | Full | Downsampled | % Female |
|---|---|---|---|---|
| ADSP | AD | 4697 Cases | 955 Cases | 60.31 |
| | | 4317 Controls | | |
| ADNI | AD | 45 Cases | 13 Cases | 38.46 |
| | | 254 Controls | | |
| Joint Genotyping[a] | AD | 806 Cases | 32 Cases | 40.63 |
| | | 908 Controls | | |
| AMP-PD | PD | 2470 Cases | 1000 Cases | 39.90 |
| | | 2023 Controls | | |
| DementiaSeq | ALS | 1105 Cases | 1000 Cases | 45.10 |
| | LBD | 2603 Cases | 1000 Cases | 34.10 |
| | FTD | 1464 Cases | 1000 Cases | 46.30 |
| | | 3193 Controls[b] | | |

[a]Joint genotyping is made up of MayoRNAseq, MSBB, and ROSMAP.
[b]Controls were shared across diseases (ALS. LBD, and FTD) in the DementiaSeq cohort.

**Supplementary Table 2 Quality control metrics by cohort**

| | **ADSP** | **ADNI** | **MayoRNAseq/ROSMAP /MSBB** | **AMP-PD** | **DementiaSeq** |
|---|---|---|---|---|---|
| **Individual QC Step** | **Number of Individuals** | | | | |
| Genotype Missingness Prune | 442 | 0 | 0 | 0 | 0 |
| Genetic Sex Confirmation | NA | 0 | 12 | 0 | NA |
| Related Prune | 2923 | 15 | 58 | 41 | 0 |
| Total Pruned | 3365 | 15 | 70 | 41 | 0 |
| Total Remaining | 9014 | 299 | 1714 | 4493 | 8365 |
| **Variant QC Step** | **Number of SNPs** | | | | |
| Callrate Prune | 14,054,936 | 2,969,411 | 683,372 | 2,931,688 | 0 |
| Case/Control Missingness Prune | 386,306 | 47 | 159 | 300,496 | 0 |
| Haplotype Prune | 1,426,302 | 138,303 | 130,016 | 235,843 | 0 |
| Hardy-Weinberg Equilibrium Prune | 578,638 | 28,701 | 480,144 | 698,633 | 3824 |
| Total Pruned | 16,446,182 | 3,136,462 | 1,293,691 | 4,166,660 | 3824 |
| Total Remaining | 206,582,127 | 41,331,241 | 58,359,334 | 168,929,789 | 215,514,196 |

More detailed information on each QC step can be found in the GP2 pipeline descriptions (https://github.com/GP2code/).

**Supplementary Table 3 Quality control metrics for merged data**

| QC Step | Number of Individuals |
|---|---|
| Related Prune | 3929 |
| Ancestry Prune | 4325 |
| Total Remaining | 16,030 |

More detailed information on each QC step can be found in the GP2 pipeline descriptions (https://github.com/GP2code/).

**Supplementary Table 4 Average number of samples in cluster 0 across the different 70:30 (training:testing) splits in the multi-disease iterative clustering analysis**

| Training:Testing Split | Average number of samples in Cluster 0 |
|---|---|
| 1 | 4571.46 |
| 2 | 4329.87 |
| 3 | 4344.95 |
| 4 | 4382.14 |
| 5 | 4430.58 |
| 6 | 4360.83 |
| 7 | 4322.33 |
| 8 | 4509.59 |
| 9 | 4307.75 |
| 10 | 4090.93 |
| 11 | 4419.70 |
| 12 | 4645.17 |
| 13 | 4305.07 |
| 14 | 4603.89 |
| 15 | 4412.52 |

**Supplementary Table 5 PheWAS for specified variants important in determining multi-disease cluster membership**

| Variant | Train | Beta | P-value |
|---|---|---|---|
| rs72654445_AD_APOC1 | Low density lipoprotein cholesterol levels | 0.082 | 3.90E-22 |
| | AD or family history of AD | -0.272 | 5.80E-09 |
| | Lymphocyte counts | -0.039 | 1.20E-05 |
| | Liver enzyme levels (alanine transaminase) | 7.20E-03 | 2.10E-05 |
| | Monocyte count | -0.030 | 1.20E-04 |
| rs112952132_AD_CEACAM16_AS1 | Low density lipoprotein cholesterol levels | -0.268 | 3.30E-168 |
| | AD or family history of AD | -0.404 | 2.10E-15 |
| | High cholesterol | -0.240 | 6.20E-11 |
| | Red cell distribution width | 0.058 | 5.20E-09 |
| | Mean spheric corpuscular volume | 0.052 | 1.50E-06 |
| rs111278137_AD_CEACAM16_AS1 | Low density lipoprotein cholesterol levels | -0.113 | 1.00E-65 |
| | AD or family history of AD | -0.299 | 4.50E-15 |
| | High cholesterol | -0.014 | 8.40E-09 |
| | Red cell distribution width | -0.116 | 5.90E-05 |
| | Glycine levels | 0.125 | 2.10E-04 |
| rs41290102_AD_NECTIN2 | Triglyceride levels | -8.60E-02 | 5.20E-25 |
| | C-reactive protein levels | 0.072 | 4.00E-19 |
| | Mean platelet volume | 0.0548 | 3.50E-16 |
| | AD or family history of AD | -0.277 | 7.00E-13 |
| | Serum alkaline phosphatase levels | 0.040 | 4.70E-07 |
| rs79701229_AD_NECTIN2 | Low density lipoprotein cholesterol levels | 0.188 | 1.90E-118 |
| | AD or family history of AD | 1.070 | 2.50E-109 |
| | High cholesterol | 0.020 | 6.70E-12 |
| | Cholesterol lowering medication | 0.235 | 1.10E-08 |
| | Platelet distribution width | 0.049 | 1.30E-07 |

**Supplementary Table 6 Top variants in determining multi-disease cluster memberships that are consistent across clustering iterations**

| Cluster 0 | Cluster 1 | Cluster 2 |
|---|---|---|
| rs72654445_AD_APOC1 | rs72654445_AD_APOC1 | rs112952132_AD_CEACAM16_AS1 |
| rs41290102_AD_NECTIN2 | rs41290102_AD_NECTIN2 | rs200046586_AD_GEMIN7 |
| rs117261169_AD_CLPTM1 | rs117261169_AD_CLPTM1 | rs111278137_AD_CEACAM16_AS1 |
| rs114088559_PD_LOC105377329 | rs114088559_PD_LOC105377329 | rs2965163_AD_GEMIN7 |
| rs112450640_AD_CBLC | rs8102895_AD_CLPTM1 | rs80307900_AD_GEMIN7 |
| rs8102895_AD_CLPTM1 | rs12983572_AD_CLPTM1 | rs713522_PD_MAPT |
| rs79701229_AD_NECTIN2 | rs79701229_AD_NECTIN2 | rs2965109_AD_CEACAM16_AS1 |
| rs12983572_AD_CLPTM1 | rs183442275_AD_MARK4 | rs35385129_AD_ZNF233 |
| rs183442275_AD_MARK4 | rs6599388_PD_LBD_TMEM175 | rs11767557_AD_EPHA1_AS1 |
| rs75627662_AD_MARK4 | rs75627662_AD_MARK4 | |
| rs112952132_AD_CEACAM16_AS1 | rs148933445_AD_CBLC | |
| rs139185008_ALS_MOB3B | rs139185008_ALS_MOB3B | |
| rs12691088_AD_LBD_APOC1 | rs41290102_AD_NECTIN2 | |

**Supplementary Table 7 SNP association summary statistic per multi-disease cluster for variants associated with loci with previously establish pleiotropic associations (*GBA, GRN, LRRK2, MAPT, APOE, C9orf72)***

| Cluster | SNP | BETA | SE | OR | P |
|---|---|---|---|---|---|
| Cluster 0 | rs17696570_ALS_C9orf72 | -0.222 | 0.035 | 0.801 | 2.74E-10 |
| | rs537741299_LBD_APOE | -5.346 | 1.216 | 0.005 | 1.10E-05 |
| | rs28903073_PD_LRRK2 | -0.397 | 0.111 | 0.672 | 3.60E-04 |
| | rs769446_AD_APOE | 0.193 | 0.079 | 1.213 | 0.015 |
| | rs713522_PD_MAPT | 0.075 | 0.034 | 1.078 | 0.029 |
| | rs76763715_PD_GBA | -1.618 | 0.777 | 0.198 | 0.037 |
| | rs10812619_ALS_C9orf72 | 0.069 | 0.035 | 1.072 | 0.045 |
| Cluster 1 | rs17696570_ALS_C9orf72 | 0.229 | 0.035 | 1.257 | 6.07E-11 |
| | rs28903073_PD_LRRK2 | 0.272 | 0.061 | 1.312 | 7.88E-06 |
| | rs537741299_LBD_APOE | 4.670 | 1.220 | 106.751 | 1.29E-04 |
| | rs76763715_PD_GBA | 2.082 | 0.782 | 8.020 | 0.008 |
| | rs713522_PD_MAPT | -0.089 | 0.035 | 0.914 | 0.010 |
| | rs769446_AD_APOE | -0.191 | 0.076 | 0.826 | 0.012 |
| Cluster 2 | rs537741299_LBD_APOE | 11.713 | 4.907 | 122210.47 | 0.017 |
| | rs76763715_PD_GBA | -7.091 | 3.110 | 8.33E-04 | 0.023 |
| | rs28903073_PD_LRRK2 | 0.138 | 0.072 | 1.148 | 0.054 |

**Supplementary Table 8 *APOE* rs7412 association summary statistic per multi-disease cluster**

| **Cluster** | **BETA** | **SE** | **OR** | **P** |
|---|---|---|---|---|
| Cluster 0 | 0.045 | 0.044 | 1.047 | 0.296 |
| Cluster 1 | -0.047 | 0.043 | 0.954 | 0.275 |
| Cluster 2 | 0.065 | 0.360 | 1.067 | 0.857 |

**Supplementary Table 9 PRS mean and standard deviation summary statistics per disease-specific cluster and counts for disease-specific cluster membership**

| Disease | Cluster | AD PRS | PD PRS | ALS PRS | LBD PRS | FTD PRS | # samples in cluster |
|---|---|---|---|---|---|---|---|
| AD | Cluster 0 | 8.00e-03 (0.951) | -0.019 (0.957) | -0.084 (0.895)* | -0.058 (0.979) | -0.035 (1.021) | 622 |
| | Cluster 1 | 0.145 (1.037)* | -0.048 (1.058) | 0.231 (1.165)* | 0.213 (1.010)* | 0.061 (0.971) | 364 |
| | Cluster 2 | -0.397 (0.771) | 0.165 (0.761) | 0.107 (1.345) | -0.418 (0.757) | -0.292 (0.904) | 12 |
| PD | Cluster 0 | 0.095 (0.986)* | -0.036 (1.008) | -0.034 (0.925) | -0.031 (1.076) | 0.017 (1.004) | 643 |
| | Cluster 1 | -0.059 (1.094) | 0.096 (1.115) | 0.024 (1.090) | 0.137 (0.975)* | 5.01e-03 (1.010) | 357 |
| ALS | Cluster 0 | 0.055 (0.999) | -0.057 (0.948) | -0.123 (0.901)* | -0.033 (0.990) | -0.006 (1.021) | 818 |
| | Cluster 1 | -0.150 (0.981)* | 0.172 (1.087)* | 0.422 (1.246)* | 0.134 (1.001) | 0.073 (0.969) | 182 |
| LBD | Cluster 0 | -0.159 (0.867)* | -0.041 (0.931) | -0.034 (0.945) | -0.226 (0.927)* | -0.081 (1.032) | 534 |
| | Cluster 1 | 0.113 (1.099)* | 0.044 (1.045) | 0.079 (1.116) | 0.250 (1.021)* | 0.070 (0.958) | 466 |
| FTD | Cluster 0 | -0.039 (0.868) | -0.113 (0.915)* | -0.114 (0.826)* | -0.129 (0.971)* | 0.013 (0.974) | 611 |
| | Cluster 1 | 0.130 (1.151)* | 0.159 (1.114)* | 0.182 (1.149)* | 0.174 (1.086)* | 0.013 (0.986) | 364 |
| | Cluster 2 | -1.105 (0.789)* | 0.028 (0.904) | 0.009 (1.235) | -0.234 (0.949) | -0.081 (1.266) | 25 |

Format: mean (standard deviation).
*denotes a p-value < 0.05 for the deviation of PRS from the normal distribution (mean=0, standard deviation=1) within a cluster.

**Supplementary Table 10 Multi-disease cluster counts for different approaches**

| Cluster | Iterative Clustering | Hierarchical Clustering | Downsampled | Controls |
|---|---|---|---|---|
| Cluster 0 | 2863 | 4073 | 1464 | 3743 |
| Cluster 1 | 2074 | 857 | 1006 | 2222 |
| Cluster 2 | 63 | 70 | 30 | 35 |
| Total | 5000 | 5000 | 2500 | 6000 |

**Supplementary Table 11 Disease enrichments per multi-disease hierarchical cluster**

| Disease | Multi-disease cluster membership | % samples with disease |
|---|---|---|
| AD | Cluster 0 | 0.177* |
| | Cluster 1 | 0.320* |
| | Cluster 2 | 0.071* |
| PD | Cluster 0 | 0.205 |
| | Cluster 1 | 0.174* |
| | Cluster 2 | 0.228* |
| ALS | Cluster 0 | 0.213 |
| | Cluster 1 | 0.146* |
| | Cluster 2 | 0.114* |
| LBD | Cluster 0 | 0.198 |
| | Cluster 1 | 0.210 |
| | Cluster 2 | 0.200 |
| FTD | Cluster 0 | 0.207 |
| | Cluster 1 | 0.150* |
| | Cluster 2 | 0.386* |

**Supplementary Table 12 Disease association summary statistics and frequency per multi-disease cluster with the *APOE* locus removed**

| Disease | Multi-disease cluster membership | OR | BETA | SE | P | % samples with disease |
|---|---|---|---|---|---|---|
| AD | Cluster 0 | 0.619 | -0.479 | 0.086 | 2.31E-08 | 0.163* |
| | Cluster 1 | 1.701 | 0.531 | 0.086 | 6.12E-10 | 0.254* |
| | Cluster 2 | 0.415 | -0.879 | 0.431 | 0.041 | 0.090* |
| PD | Cluster 0 | 1.117 | 0.111 | 0.086 | 0.196 | 0.208 |
| | Cluster 1 | 0.907 | -0.098 | 0.086 | 0.258 | 0.191 |
| | Cluster 2 | 0.813 | -0.207 | 0.334 | 0.536 | 0.157* |
| ALS | Cluster 0 | 1.752 | 0.561 | 0.090 | 4.76E-10 | 0.229* |
| | Cluster 1 | 0.587 | -0.533 | 0.091 | 4.48E-09 | 0.160* |
| | Cluster 2 | 0.574 | -0.553 | 0.380 | 0.145 | 0.202 |
| LBD | Cluster 0 | 0.838 | -0.177 | 0.084 | 0.035 | 0.191 |
| | Cluster 1 | 1.132 | 0.125 | 0.084 | 0.139 | 0.210 |
| | Cluster 2 | 1.631 | 0.489 | 0.276 | 0.076 | 0.270* |
| FTD | Cluster 0 | 1.016 | 0.016 | 0.085 | 0.851 | 0.209 |
| | Cluster 1 | 0.943 | -0.058 | 0.085 | 0.497 | 0.184* |
| | Cluster 2 | 1.855 | 0.618 | 0.269 | 0.021 | 0.281* |

*denotes a p-value $< 0.05$ for the frequency increase or decrease in a certain disease status per cluster compared to the null estimate of 20%.

**Supplementary Table 13 PRS association summary statistics per multi-disease cluster with the *APOE* locus removed**

| Disease | Cluster | OR | BETA | SE | P |
|---|---|---|---|---|---|
| AD | Cluster 0 | 0.771 | -0.260 | 0.045 | 8.37E-09 |
| | Cluster 1 | 1.330 | 0.284 | 0.045 | 4.00E-10 |
| | Cluster 2 | 0.743 | -0.296 | 0.180 | 0.101 |
| PD | Cluster 0 | 0.819 | -0.199 | 0.045 | 1.20E-05 |
| | Cluster 1 | 1.229 | 0.206 | 0.046 | 7.00E-06 |
| | Cluster 2 | 0.948 | -0.053 | 0.161 | 0.743 |
| ALS | Cluster 0 | 0.852 | -0.160 | 0.045 | 3.25E-04 |
| | Cluster 1 | 0.178 | 0.163 | 0.045 | 2.61E-04 |
| | Cluster 2 | 0.984 | -0.016 | 0.161 | 0.918 |
| LBD | Cluster 0 | 0.643 | -0.442 | 0.047 | 4.94E-21 |
| | Cluster 1 | 1.586 | 0.461 | 0.047 | 1.47E-22 |
| | Cluster 2 | 0.816 | -2.03E-01 | 0.177 | 0.251 |
| FTD | Cluster 0 | 1.009 | 0.01 | 0.045 | 0.827 |
| | Cluster 1 | 0.989 | -0.011 | 0.045 | 0.814 |
| | Cluster 2 | 1.008 | 0.009 | 0.16 | 0.956 |

**Supplementary Table 14 Disease association summary statistics and frequency per downsampled multi-disease cluster (500 cases per disease)**

| Disease | Multi-disease cluster membership | OR | BETA | SE | P | % samples with disease |
|---|---|---|---|---|---|---|
| AD | Cluster 0 | 0.594 | -0.521 | 0.121 | 1.80E-05 | 0.162* |
| | Cluster 1 | 1.735 | 0.551 | 0.121 | 6.00E-06 | 0.258* |
| | Cluster 2 | 0.557 | -0.584 | 0.618 | 0.345 | 0.100* |
| PD | Cluster 0 | 1.129 | 0.121 | 0.121 | 0.316 | 0.204 |
| | Cluster 1 | 0.886 | -0.121 | 0.122 | 0.320 | 0.194 |
| | Cluster 2 | 0.972 | -0.029 | 0.504 | 0.954 | 0.233* |
| ALS | Cluster 0 | 1.731 | 0.549 | 0.127 | 1.50E-05 | 0.232* |
| | Cluster 1 | 0.592 | -0.525 | 0.128 | 3.90E-05 | 0.157* |
| | Cluster 2 | 0.541 | -0.613 | 0.618 | 0.321 | 0.100* |
| LBD | Cluster 0 | 0.846 | -0.167 | 0.119 | 0.162 | 0.193 |
| | Cluster 1 | 1.185 | 0.170 | 0.120 | 0.156 | 0.210 |
| | Cluster 2 | 0.972 | -0.029 | 0.504 | 0.954 | 0.200 |
| FTD | Cluster 0 | 1.040 | 0.039 | 0.122 | 0.747 | 0.210 |
| | Cluster 1 | 0.903 | -0.103 | 0.124 | 0.407 | 0.181 |
| | Cluster 2 | 2.351 | 0.855 | 0.421 | 0.042 | 0.367* |

*denotes a p-value < 0.05 for the frequency increase or decrease in a certain disease status per cluster compared to the null estimate of 20%.

**Supplementary Table 15 PRS association summary statistics per downsampled multi-disease cluster**

| Disease | Cluster | OR | BETA | SE | P |
|---|---|---|---|---|---|
| AD | Cluster 0 | 0.804 | -0.218 | 0.064 | 6.78E-04 |
| | Cluster 1 | 1.317 | 0.275 | 0.065 | 2.10E-05 |
| | Cluster 2 | 0.149 | -1.902 | 0.505 | 1.66E-04 |
| PD | Cluster 0 | 0.867 | -0.143 | 0.065 | 0.027 |
| | Cluster 1 | 1.164 | 0.152 | 0.065 | 0.019 |
| | Cluster 2 | 0.867 | -0.142 | 0.286 | 0.619 |
| ALS | Cluster 0 | 0.824 | -0.193 | 0.064 | 2.50E-03 |
| | Cluster 1 | 1.229 | -0.206 | 0.064 | 1.33E-03 |
| | Cluster 2 | 0.793 | -0.232 | 0.314 | 0.459 |
| LBD | Cluster 0 | 0.678 | -0.389 | 0.066 | 3.63E-09 |
| | Cluster 1 | 1.484 | 0.395 | 0.066 | 2.39E-09 |
| | Cluster 2 | 0.955 | -0.046 | 0.286 | 0.872 |
| FTD | Cluster 0 | 0.952 | -0.048 | 0.065 | 0.456 |
| | Cluster 1 | 1.037 | 0.036 | 0.065 | 0.578 |
| | Cluster 2 | 1.323 | 0.280 | 0.335 | 0.404 |

**Supplementary Table 16 Disease association summary statistics and frequency per multi-disease cluster with controls included**

| Disease | Multi-disease cluster membership | OR | BETA | SE | P | % samples with disease |
|---|---|---|---|---|---|---|
| Control | Cluster 0 | 108.69 | 4.688 | 0.450 | 2.10E-25 | 0.265* |
| | Cluster 1 | 9.47E-03 | -4.660 | 0.450 | 4.12E-25 | 4.05E-03* |
| | Cluster 2 | 1.08E-08 | -18.340 | 4353.967 | 0.997 | 0* |
| AD | Cluster 0 | 0.485 | -0.723 | 0.084 | 8.76E-18 | 0.123* |
| | Cluster 1 | 2.097 | 0.740 | 0.084 | 1.49E-18 | 0.241* |
| | Cluster 2 | 0.442 | -0.816 | 0.738 | 0.269 | 0.114* |
| PD | Cluster 0 | 0.748 | -0.29 | 0.084 | 5.85E-04 | 0.151* |
| | Cluster 1 | 1.302 | 0.264 | 0.085 | 1.82E-03 | 0.191* |
| | Cluster 2 | 2.398 | 0.874 | 0.431 | 0.042 | 0.286* |
| ALS | Cluster 0 | 1.097 | 0.092 | 0.085 | 0.278 | 0.170 |
| | Cluster 1 | 0.927 | -0.076 | 0.085 | 0.372 | 0.162 |
| | Cluster 2 | 0.415 | -0.879 | 0.738 | 0.234 | 0.086* |
| LBD | Cluster 0 | 0.591 | -0.525 | 0.082 | 1.79E-10 | 0.139* |
| | Cluster 1 | 1.710 | 0.537 | 0.082 | 7.62E-11 | 0.214* |
| | Cluster 2 | 0.652 | -0.427 | 0.617 | 0.488 | 0.143* |
| FTD | Cluster 0 | 0.719 | -0.331 | 0.084 | 8.99E-05 | 0.152* |
| | Cluster 1 | 1.336 | 0.290 | 0.085 | 6.19E-04 | 0.188* |
| | Cluster 2 | 3.432 | 1.233 | 0.410 | 2.65E-03 | 0.371* |

*denotes a p-value < 0.05 for the frequency increase or decrease in a certain disease status per cluster compared to the null estimate of 20%.

**Supplementary Table 17 PRS association summary statistics per multi-disease cluster with controls included**

| Disease | Cluster | OR | BETA | SE | P |
|---|---|---|---|---|---|
| AD | Cluster 0 | 0.740 | -0.301 | 0.040 | 9.85E-14 |
| | Cluster 1 | 1.393 | 0.331 | 0.041 | 4.03E-16 |
| | Cluster 2 | 0.144 | -1.935 | 0.426 | 5.45E-06 |
| PD | Cluster 0 | 0.813 | -0.206 | 0.041 | 5.16E-07 |
| | Cluster 1 | 1.233 | 0.209 | 0.041 | 4.11E-07 |
| | Cluster 2 | 0.968 | -0.032 | 0.253 | 0.898 |
| ALS | Cluster 0 | 0.850 | -0.163 | 0.040 | 4.70E-05 |
| | Cluster 1 | 1.191 | 0.175 | 0.040 | 1.4E-05 |
| | Cluster 2 | 0.592 | -0.524 | 0.327 | 0.108 |
| LBD | Cluster 0 | 0.601 | -0.508 | 0.042 | 6.51E-34 |
| | Cluster 1 | 1.669 | 0.512 | 0.042 | 3.36E-34 |
| | Cluster 2 | 0.993 | -6.50E-03 | 0.252 | 0.979 |
| FTD | Cluster 0 | 0.981 | -0.019 | 0.041 | 0.636 |
| | Cluster 1 | 1.031 | 0.030 | 0.041 | 0.466 |
| | Cluster 2 | 0.722 | -0.326 | 0.213 | 0.125 |

**Supplementary Table 18 LDSC genetic correlation results between diseases**

| Disease | AD | PD | ALS | LBD | FTD |
|---|---|---|---|---|---|
| **AD** | x | 0.197 (0.084)* | 0.154 (0.120) | 0.385 (0.188)* | 0.311 (0.271) |
| **PD** | 0.197 (0.084)* | x | 0.057 (0.087) | 0.599 (0.166)* | 0.464 (0.275) |
| **ALS** | 0.154 (0.120) | 0.057 (0.087) | x | 0.121 (0.220) | 0.168 (0.324) |
| **LBD** | 0.385 (0.188)* | 0.599 (0.166)* | 0.121 (0.220) | x | 0.290 (0.515) |
| **FTD** | 0.311 (0.271) | 0.464 (0.275) | 0.168 (0.324) | 0.290 (0.515) | x |

Format: genetic correlation (standard error).
*denotes a significant correlation between diseases (p-value < 0.05).

**Supplementary Methods**

### Iterative Clustering Approach

Tracking samples across iterations allows for the variable number of clusters formed outside of the main cluster (i.e., Cluster 0) to be collapsed into Clusters 1 and 2. The variable number of clusters identified by Mean Shift is due to the stochasticity of UMAP. The iterative clustering approach is necessary to capture information provided by Mean Shift over these varying UMAP representations.

When performing the iterative clustering approach for the disease-specific analyses, the sample size was reduced from 5000 samples to 1000 samples for each NDD respectively. This reduction in sample size leads to a large increase in variability between iterations. This led to the decision to group samples into Cluster 0 or Cluster 2 (Cluster 1 if only 2 clusters were identified in a disease subset) if they were consistently inside or outside the main cluster for at least 12 of the 15 training:testing splits, instead of all 15 training:testing splits which was the approach used in the multi-disease analysis. It was necessary to reduce this threshold to account for the increased variability that occurs when performing the analysis on a significantly smaller sample size.
